# Population Pharmacokinetics of Oxcarbazepine: A Systematic Review

**DOI:** 10.1101/2021.01.27.21249807

**Authors:** Yue-ting Chen, Chen-yu Wang, Yi-wei Yin, Zi-ran Li, Wei-wei Lin, Min Zhu, Zheng Jiao

## Abstract

**Introduction:** Oxcarbazepine is commonly used as a first-line drug in the treatment of partial seizures. Due to the high pharmacokinetic variability of oxcarbazepine, many population pharmacokinetic models have been developed to optimise the dosing regimen of oxcarbazepine.

**Areas covered:** This review summarize the published population pharmacokinetic studies of oxcarbazepine in children and adults. The quality of the identified reports from the PubMed and Embase databases was also evaluated. We also explored the significant covariates that may have an impact on the dosage regimen and clinical use of oxcarbazepine.

**Expert Opinion:** The oxcarbazepine dose regimen was dependent on weight and co-administration with enzyme-inducing medications. In order to achieve more accurate treatment, we should establish PK / PD model of OXC to evaluate the effectiveness of dose adjustment from pharmacodynamic indicators. Furthermore, exploring the pharmacokinetic in specifical patients, such as infants is essential to improve its safety.

**Article highlights:**

- In this review, we identified weight, renal function, and co-administered medications as covariates that most likely to influence oxcarbazepine pharmacokinetics.
- Comparing to adult patients, paediatric patients show a higher clearance per kilogramme weigh which lead to higher doses per kilogramme; they may also require therapeutic drug monitoring owing to a larger variation in clearance.
- Further studies are essential to evaluate oxcarbazepine pharmacokinetics in special populations such as infants.

## 1. Introduction

Oxcarbazepine (OXC) is a commonly used anti-epilepsy drug with a chemical structure similar carbamazepine (CBZ) [1]. It was approved by the Food and Drug Administration as a monotherapy and adjunctive therapy for partial seizures in adult and paediatric patients. It prevents seizures mainly through the blockage of voltage-dependent sodium channels, similar to CBZ [2]. Owing to its comparable effectiveness but better safety and tolerability, OXC is usually used as an alternative to CBZ in patients who are unable to tolerate CBZ [3,4].

Oxcarbazepine is completely absorbed (>95%) and quickly transformed to its active metabolite 10-hydroxycarbazepine (MHD) by cytosolic enzymes after oral administration [5,6]. Owing to its rapid metabolism, OXC has a much lower area under the concentration-time curve (AUC) than MHD *in vivo* (16.05 vs. 215.52 μg·h/ml) [7]. Thus, the effectiveness of OXC is mainly determined by measuring MHD concentration [8]. Following OXC administration, the concentration of MHD reaches a peak in approximately 2–4 h [9]. MHD has a low protein binding rate (∼39%) [10], and its volume of distribution (Vd) is between 0.3 and 0.8 L/kg [6]. MHD is excreted unchanged in the urine or eliminated in conjugation with uridine diphosphate-glucuronosyltransferase (UGT), with only a small fraction (4%) being oxidised to its dihydroxy derivative (DHD) [5,11].

In special patient populations, such as patients with renal insufficiency, patients who are co-administered OXC with enzyme-inducing antiepileptic drugs (EIAEDs) as well as the elderly and infants, the pharmacokinetics (PK) of MHD varies greatly [12]. Rouan et al. [12] reported that the mean AUC_0-168 h_ of MHD in patients with severe renal impairment was around 2–2.5 times higher than that in healthy subjects after receiving a single oral dose of OXC. A high exposure to OXC is associated with an increased incidence of side effects [13]; therefore, to ensure effectiveness and safety, therapeutic drug monitoring (TDM) is essential for patient-specific dose adjustment [14]. Ideally, individualised doses need to be developed at the beginning of treatment; however, the current TDM approach is usually implemented during the course of the treatment. The population pharmacokinetic (PPK) approach has been used to identify significant covariates that influence PK and is often used in clinical practice through Bayesian forecasting to develop individualised therapy at the beginning and even during the course of treatment [15,16].

Despite the reports of PPK study are numerous, no research has been conducted to review the PPK of MHD. Analysing and understanding the significant covariates and their relationship in different patient populations is critical for the development of appropriate regimens for individualised therapy. In this review, we aim to summarise the significant covariates affecting PK, identify unexplored covariates, and provide evidence for the model informed of OXC.

## 2. Methods

### 2.1 Search Strategy

The PubMed and Embase databases were used to systematically search for articles on PPK of OXC published before 31 October 2020 followed the principles of the Preferred Reporting Items for Systematic Reviews and Meta-analyses (PRISMA) statement. The search terms used included “oxcarbazepine”, “oxcarbamazepine”, “ocbz”, “oxtellar”, or “trileptal” and “population pharmacokinetic”, “pharmacokinetic modeling”, “nonlinear mixed effect model”, “NONMEM”, “Pmetrics”, “WINNONMIX”, “ADAPT”, “P-PHARM”, “nlmixed”, “NLME”, “USC^*^PACK”, or “MONOLIX”. Besides, references of all included studies were searched.

All the relevant articles selected from the databases and reference lists were screened to evaluate their eligibility for inclusion according to the following criteria: target group of studies was human; OXC was the study drug; PPK analysis was conducted in the study; and the study was published in English. A study was excluded if it was a review or external evaluation article; the data presented overlapped with previous studies but with smaller sample sizes; or the PK parameters were incomplete. Two independent authors screened the title and abstracts as well as the full-text of each article to determine their eligibility. Discrepancies were resolved by a third senior investigator.

### 2.2 Evaluation of Literature Quality

The quality of the PPK report was evaluated according to previous guidelines of PK reports (Kanji et al. [17] and Jamsen et al. [18]) that assist in transparent and complete reporting. A checklist with 24 items was developed to summarise the key points of the report. All items were categorised into five sections: the title and abstract, background and introduction, methods, results, and discussion and conclusion, according to their relevance to the sections of the research report.

If there was a description in the paper that conforms to the criterion, the scoring item will be counted as 1 point. If it was not reported, zero points were given. Each identified study was evaluated based on these criteria. Compliance of the included PPK studies was reported as a proportion. The compliance rate was calculated as follows:

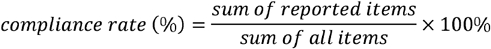

### 2.3 Data Extraction

A standardised data collection was used to extract the relevant information from each eligible study as follows: patient characteristics, such as age, weight, and laboratory test, ; the study design, such as research types, quantity of subjects and observations; dose regimens; OXC formulation; and PPK study information such as data analysis software and algorithm, model strategies, final PK parameters, and tested and significant covariates.

### 2.4 Study Comparison

We developed virtual patients with different age: infants (10 kg, 1-year-old) taking 300 mg OXC every 12 h (q12h), children (30 kg, 10 years old) taking 300 mg OXC q12h, and adults (70 kg, 40 years old) taking 600 mg OXC q12h. The concentration–time profiles in virtual patients were drew on the basis of the restored PPK model and study population in each included study. They received multiple doses of OXC alone and achieve stable state. The sex of the virtual patient was selected as male.

The identified covariates effect on clearance (CL) in the included studies was analysed and showed by a forest map. The CL change less than between 80%-125% was not considered to have a significant clinical correlation [19]. For binary covariates such as co-administration with medications, we use 0 for monotherapy and 1 for co-administration with medications. For continuous covariates that were included in only one model, we used the same range values as the study. For continuous covariates included in multiple studies, we scaled them to the same range for comparison. The effect of each covariate on CL was shown as the ratio of CL in the range of the covariate divided by the typical CL value in each study.

The simulation was conducted using NONMEM (version 7.4; ICON Development Solutions, Ellicott City, MD, USA) software. R (version 3.5.1; http://www.r-project.org/) was used to generate the concentration–time plots and forest plots.

## 3. Results

### 3.1 Study Identification

A total of 155 articles were selected: 87 articles were selected from PubMed; 67 articles from the Embase database; and one article from the references of selected articles. After initial screening, 16 full-text studies were assessed for eligibility. One study was excluded because the final PK parameters were not presented [20], and two were excluded because of language [21,22]. In addition, the study conducted by Sallas et al. [23] was excluded because the study data overlapped with that of Sugiyama et al. [24]. Finally, 12 articles were included in the study (**Fig. 1**).

**Fig. 1.**
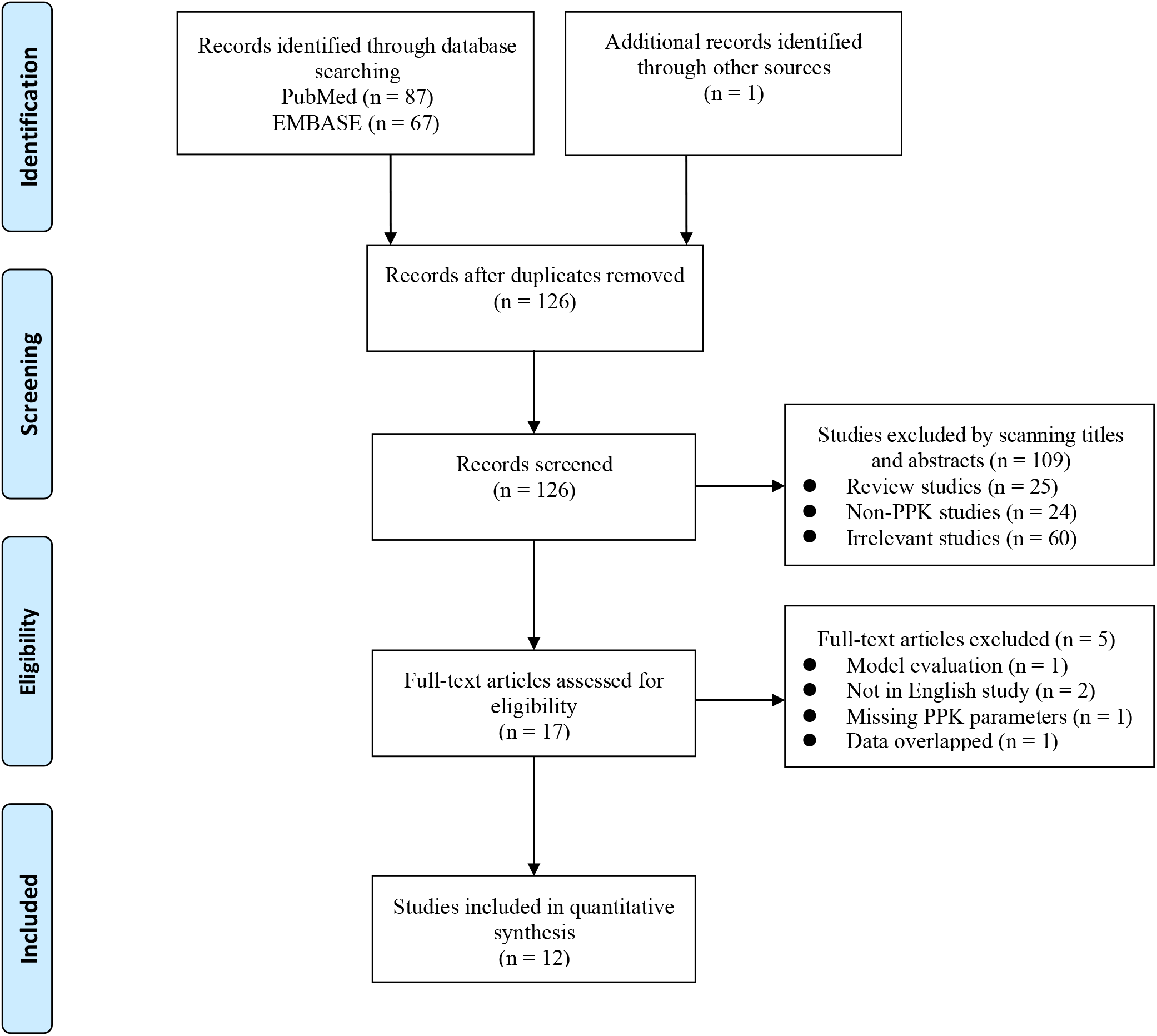
PRISMA flow diagram for the identification of population pharmacokinetic studies.

### 3.2 Evaluation of Literature Quality

According to the guidelines, we summarised the quality of OXC PPK studies based on 35 items, as outlined in **Table 1**. Compared with that of the PPK articles published between 2005 and 2013, the quality of PPK studies published between 2014 and 2019 significantly improved [compliance rate median (range): 71.4% (68.6%–71.4%) vs. 94.3% (85.7%–97.1%)]. Generally, prospective studies show better compliance than retrospective studies; however, the prospective study by Northam et al. [25] had a compliance rate of only 73.5%. This may be due to the lack of a standard reporting guide published at that time.

**Table 1.**
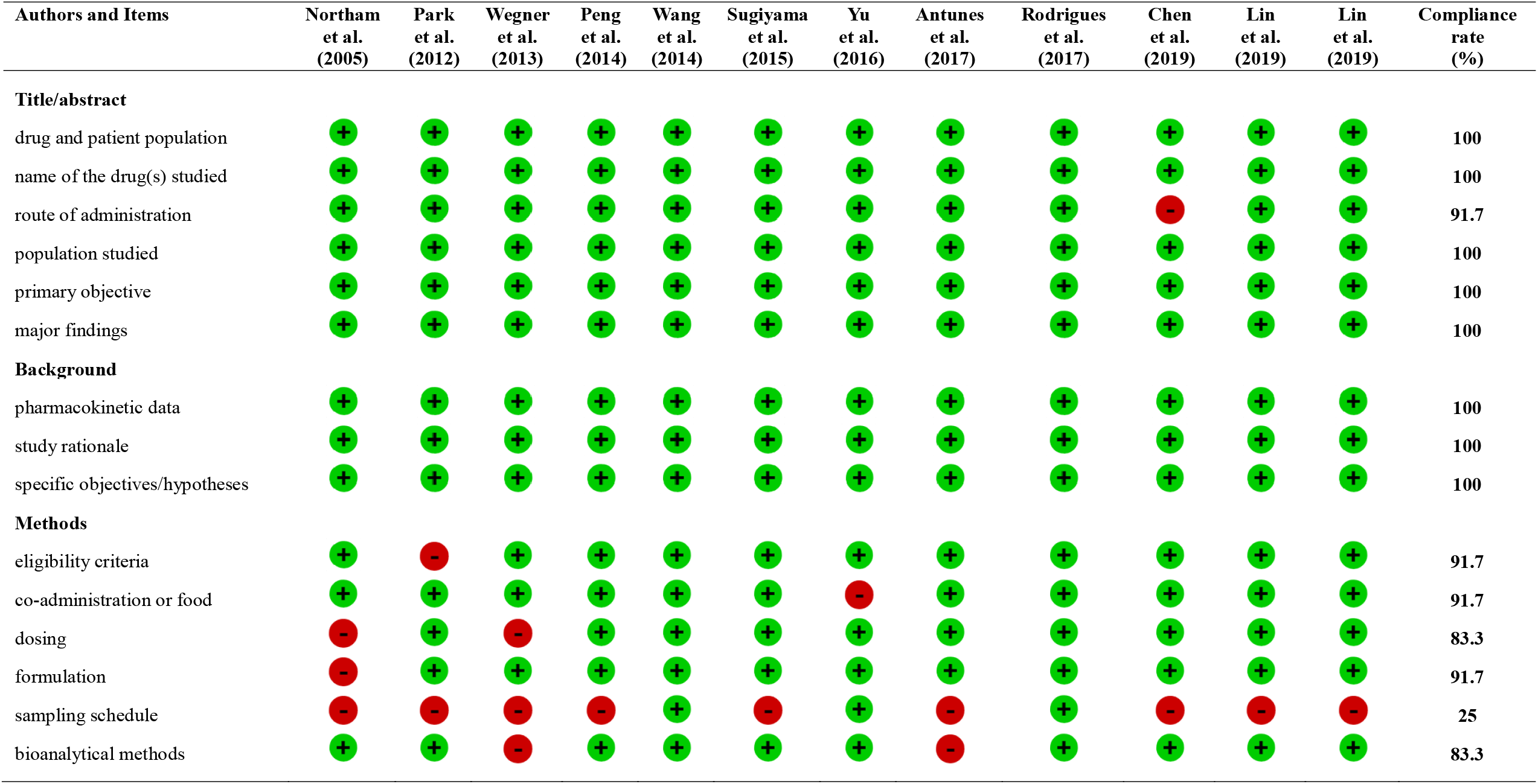
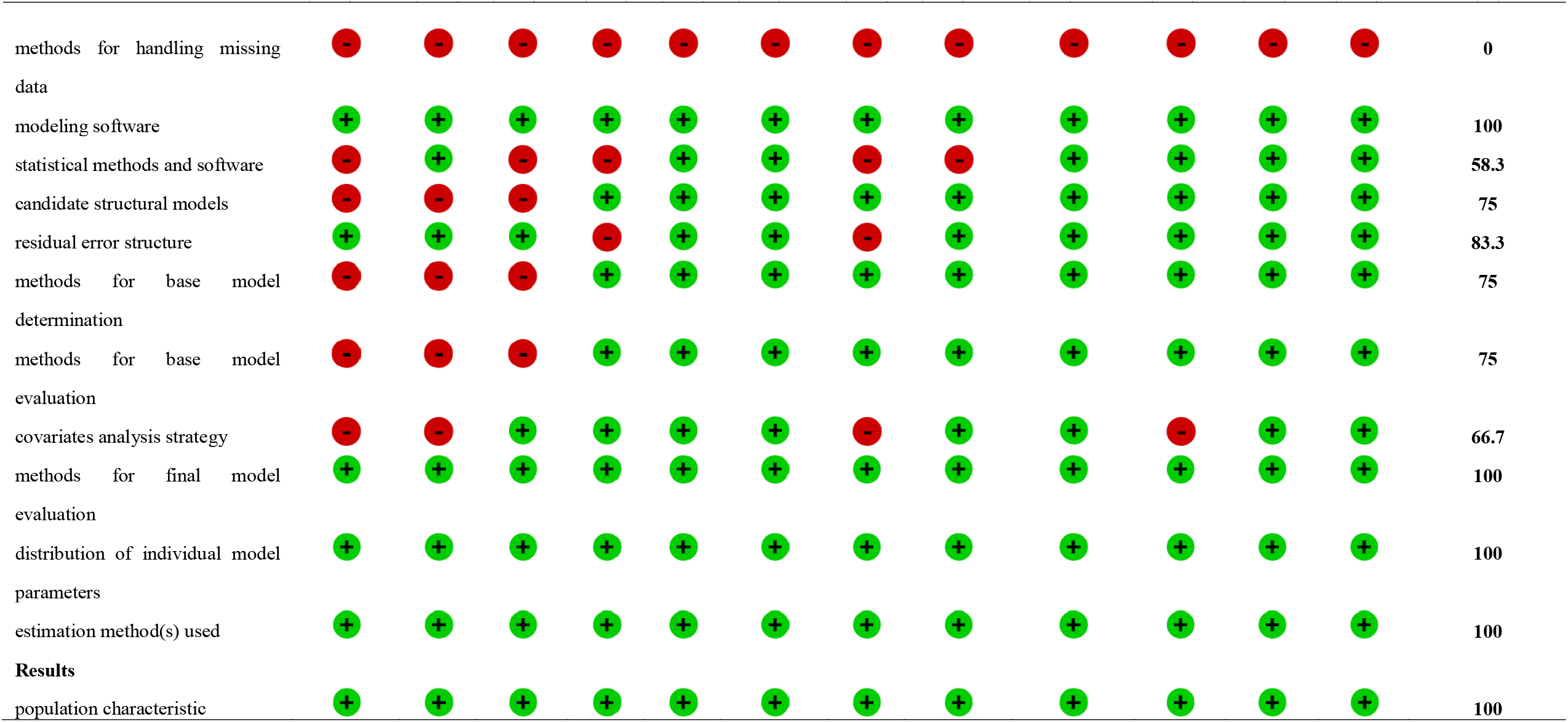
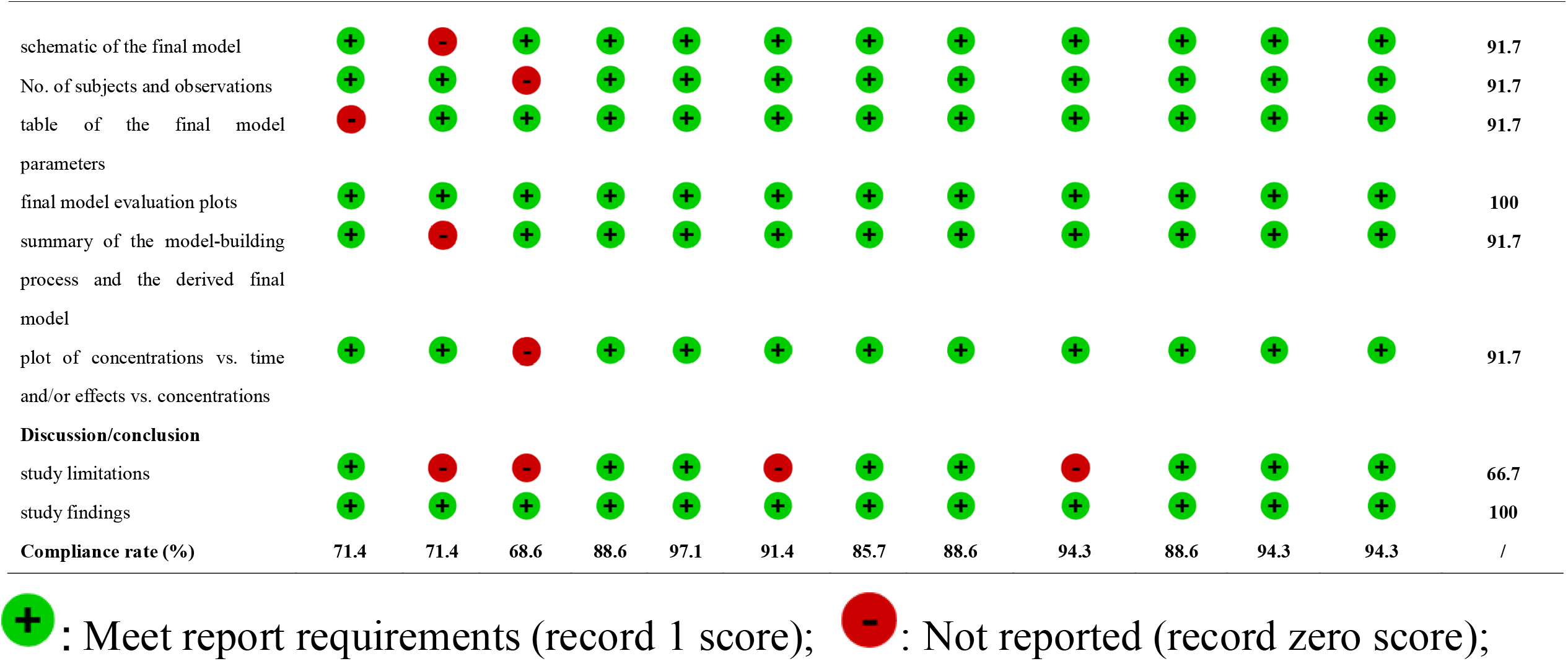
ClinPK checklist of information to be included when reporting a clinical pharmacokinetic study.

**Table 2.**
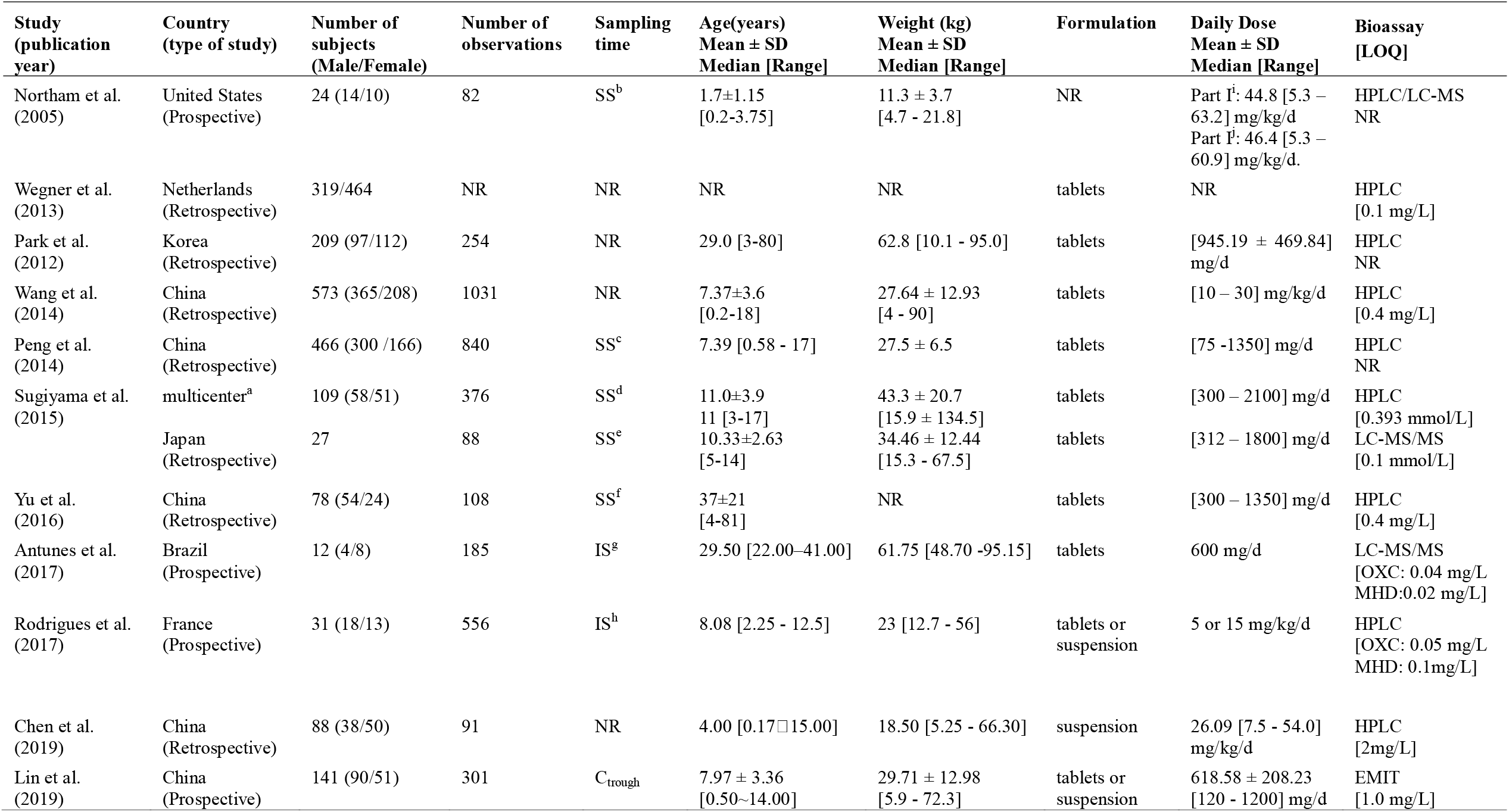
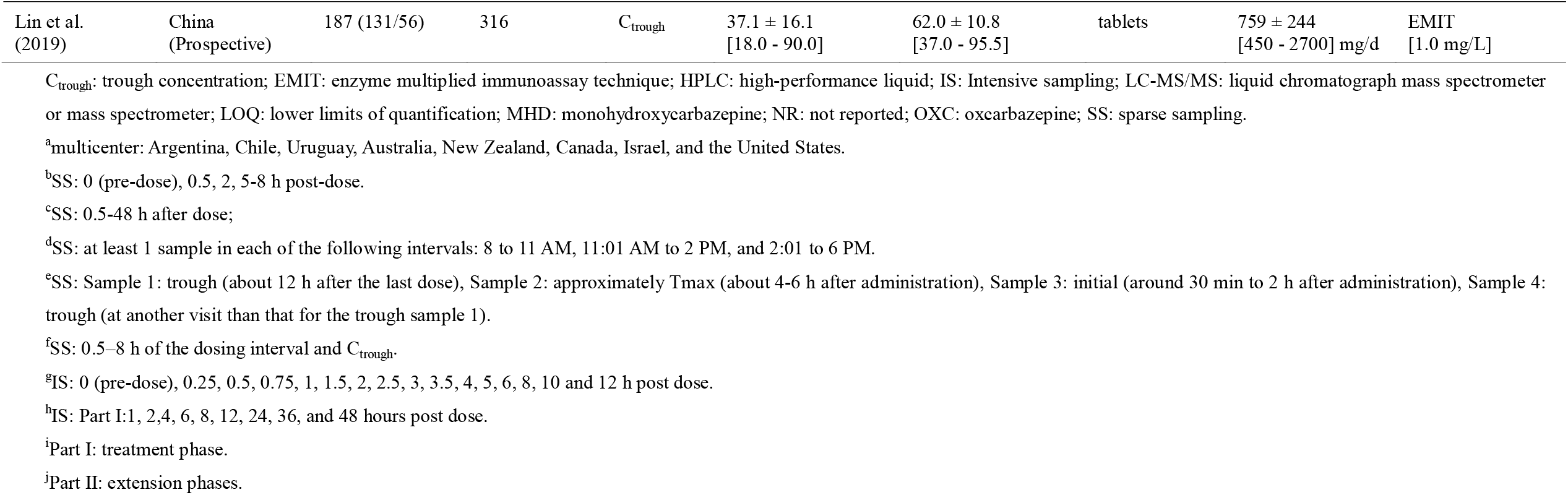
Characteristics of included population pharmacokinetic studies.

After the guidelines were published, the quality of PPK study reports improved over time. Nonetheless, several studies still have compliance rate of less than 60%, which included deficiencies in the description of sample, statistical methods and software, and methods for handling missing data.

### 3.3 Study Characteristics

The included studies were published from 2005 to 2019. The total quantity of subjects in included articles between 12 and 573. Most PPK analysis was performed in patients with epilepsy [24,26-33], and only Antunes et al. [34] enrolled healthy subjects. One study was conducted only in infants [25], seven studies were conducted in children [24,25,27,28,30-32], two studies included both children and adults [26,33], and two studies only enrolled adults [29,34]. Subjects in all the included studies were administered OXC orally, with doses ranging from 75 to 1800 mg/d. In most studies, sparse sampling was employed, with 1 to 4 samples collected per individual. Detailed characteristics of all included studies were listed in **Table 1**.

The PPK analyses were all conducted using population modelling software, including NONMEM (Icon, Dublin, Ireland) [24-26,29-31,34,35], Phoenix NLME (Certara L.P. Pharsight, St. Louis, MO, USA) [26–28], and Monolix (Lixoft, Antony, France) [32]. The most commonly used algorithm was first-order conditional estimation with the η*-*ε interaction.

Most studies described the PK of MHD as a one-compartment model with first-order absorption and elimination. Rodrigues et al. [32] described the PK of OXC and its active metabolite MHD with a two-compartment model with first-order absorption and elimination. Antunes et al. [34] described the PK of OXC and its active metabolite enantiomers using a two-compartment model with transit absorption and first-order elimination.

All studies used either internal or external evaluation. The commonly method of external evaluation is the goodness-of-fit plot, while the second popular method was the visual predictive check. Although a normalised prediction distribution error (NPDE) has better properties [36], it is not commonly used [29,30,32,34]. Four studies performed an external evaluation using an independent dataset, all of which showed acceptable predictability [28,30,31,33].

### 3.4 Study Comparison

There were no obvious differences in the PK of OXC due to ethnicity. **Fig. 2** shows concentration-time profiles of all studies that are simulated with virtual infant (a), children (b) and adult (c) patients. The model established by Peng et al. [27] which presented lower CL, displayed a much higher trough concentration (C_trough_) than others (CL: 0.016 vs. 0.035–0.073 L/h/kg). On the contrary, the PPK model established by Yu et al. [33] which presented a higher CL, showed a considerably lower C_trough_ than others (CL: 0.084 vs. 0.035–0.073 L/h/kg).

**Fig. 2.**
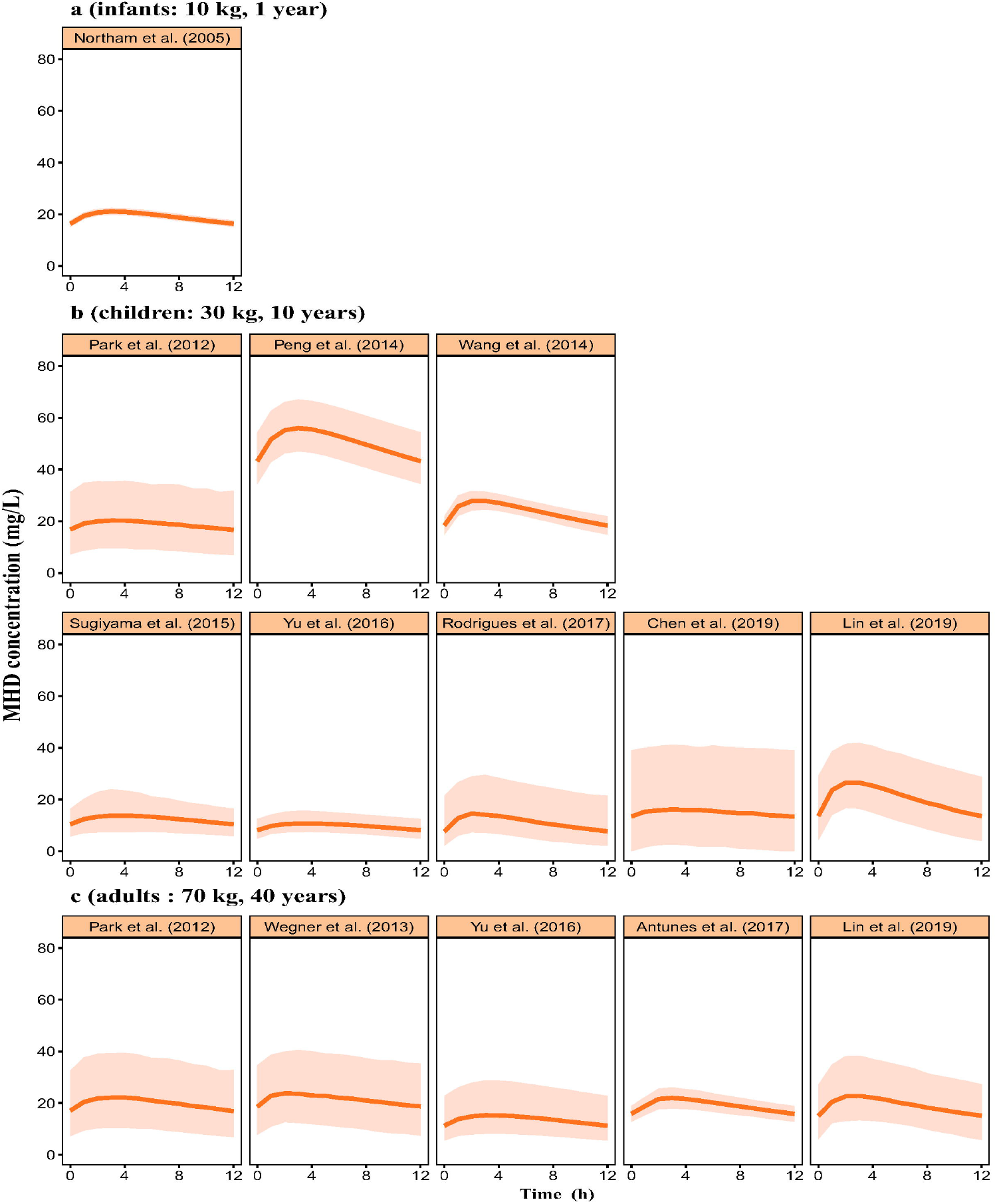
Concentration-time profiles at steady state for (a) infants, (b) children, and (c) adults in retrieved studies. The solid line represents median of the simulated concentration-time profile. The light shadows represent the 10th-90th percentiles of the simulated concentration-time profiles. The estimated glomerular filtration rate (eGFR) was set to 90 mL/min for adults. The alanine aminotransferase (ALT) for children was set to 30 U/L. Age was set to 18-75 years. All patients were assumed to be male receiving OXC monotherapy at a dose of 300 mg for infants and children; 600 mg for adults.

The estimated median CL was 0.078 L/h/kg for infants (10 kg, 1 year) [25], 0.058 (range: 0.016–0.084) L/h/kg for children (30 kg, 10 years) [24,26-28,30-33], and 0.033 (range: 0.029–0.0599) L/h/kg for adults (70 kg, 40 years) [26,29,33-35]. The median CL per kg in infants and children was higher than that in adults. Furthermore, children had a larger variability in CL than adults (0.016–0.084 L/h/kg vs. 0.029–0.06 L/h/kg).

Moreover, the median Vd per kg in infants and children was higher than that in adults. The determined median Vd was 1.38 L/kg for infants, 0.78 (range: 0.47–2.16) L/kg for children, and 0.45 (range: 0.2–1.48) L/kg for adults.

The final PK parameters included in our review are summarised in **Table 3**. The between-subject variability (BSV) was determined from exponential models in all included studies. The median (range) BSV was as follows: CL, 16.84% (11.09%–34.21%); Vd, 35.35% (6.88%–104.4%); and Ka, 24.05% (8.27%–39.82%).

**Table 3.**
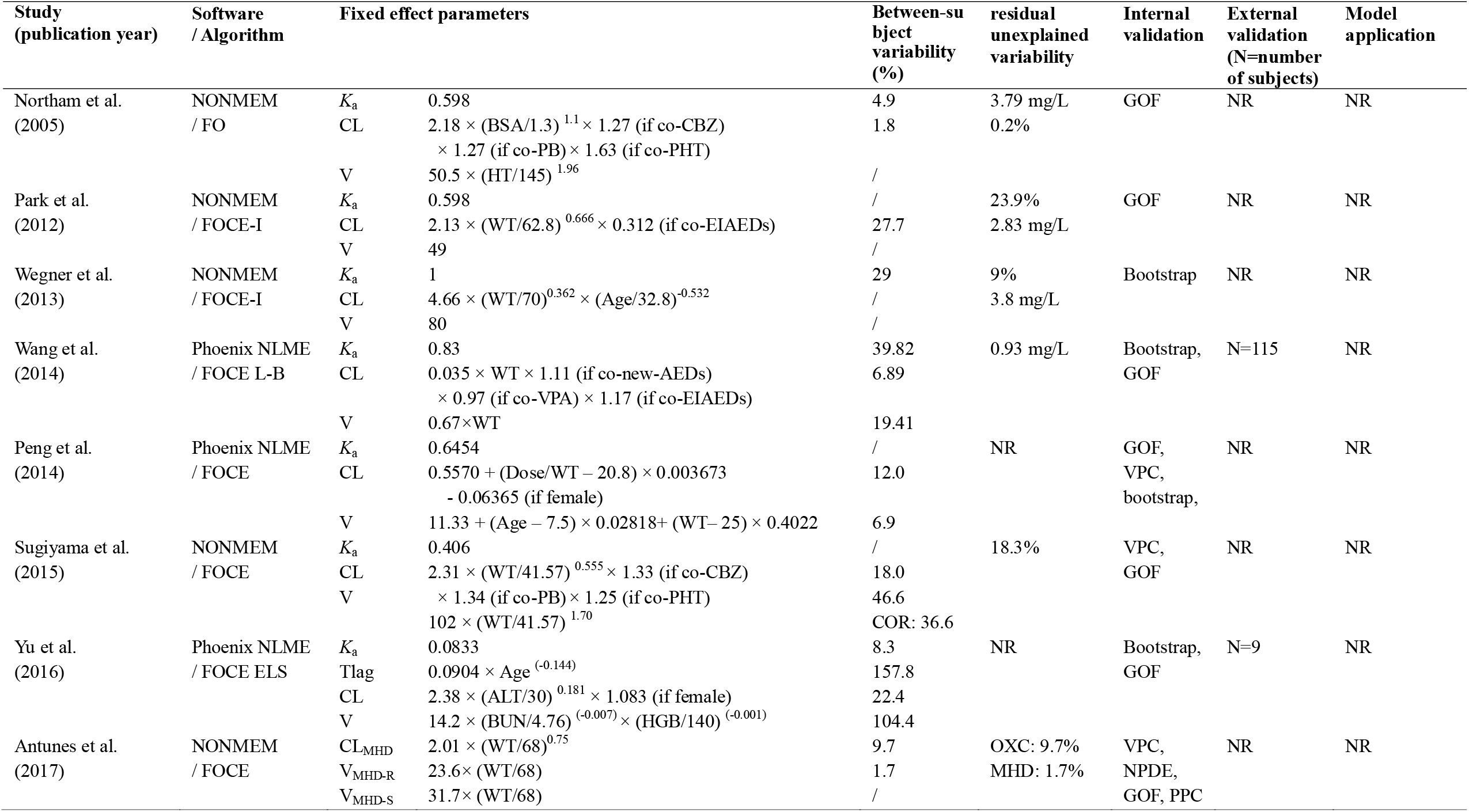
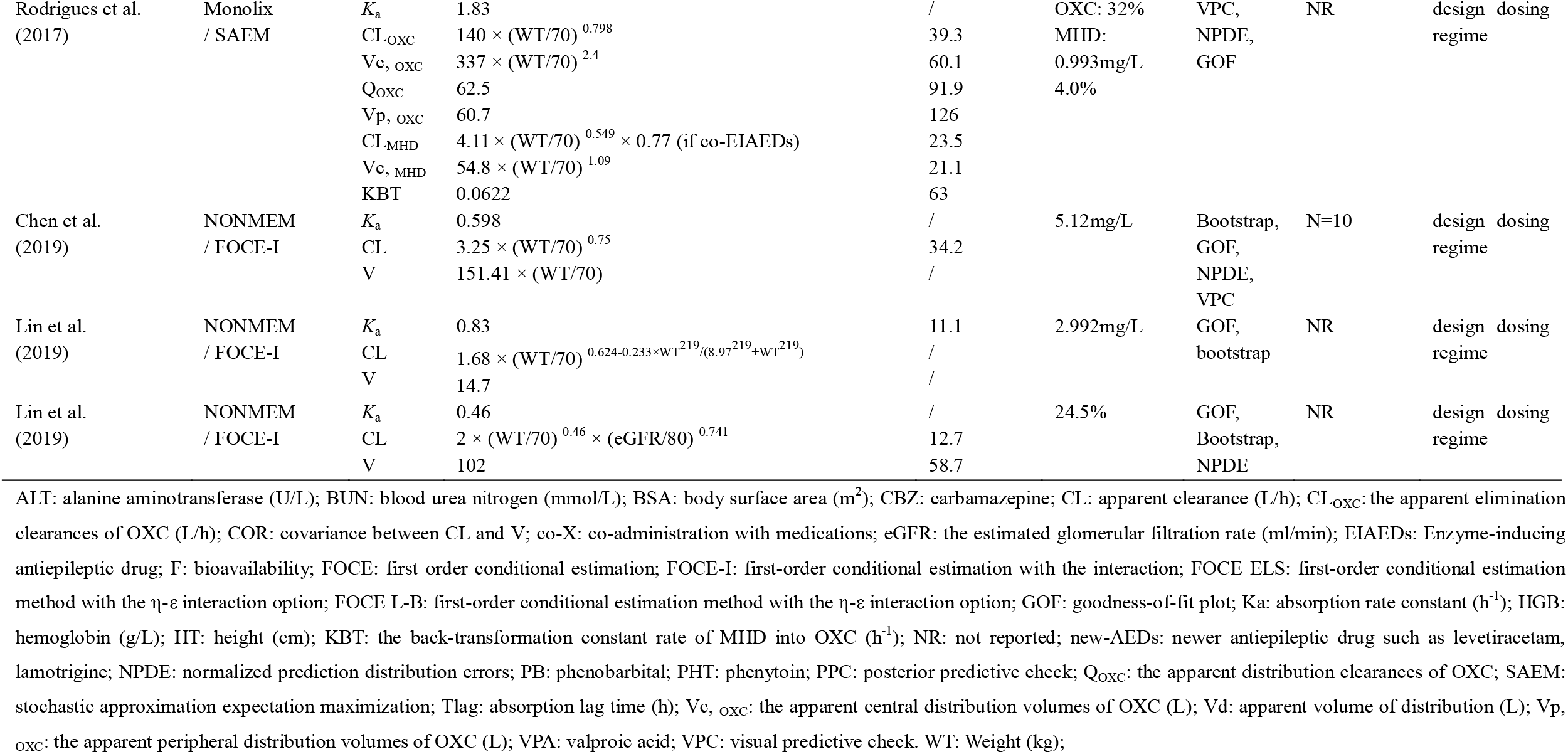
Model strategies and final pharmacokinetic parameters of included studies.

The residual unexplained variability (RUV) was described using the proportional model: 12.05% (4%–32%) or additive model: 3.396 mg/L (0.93–0.512 mg/L).

All PPK analysis intended to explain the BSV of MHD PK by investigating possible covariates. The covariates investigated and identified in each study are visually shown in **Fig. 3**. The commonly investigated covariates included weight, sex, age, co-administered medications, body surface area, and estimated glomerular filtration rate (eGFR). The identified covariates for CL included weight, co-administered medications, eGFR, dose. All the investigated and identified covariates in the PPK model are summarised in **Table S1**.

**Fig. 3.**
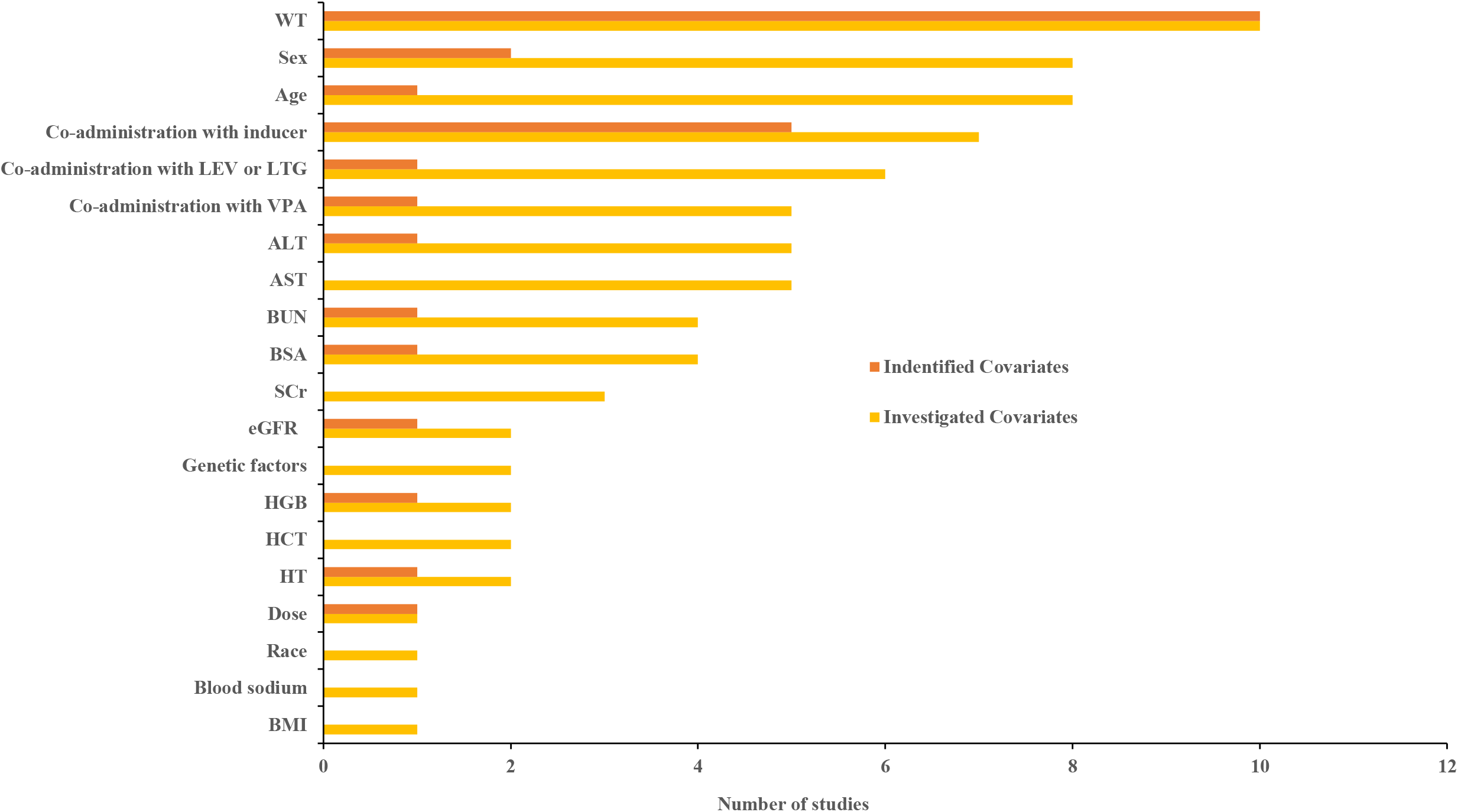
Investigated and identified covariates for clearance of oxcarbazepine. AST: aspartate aminotransferase; ALT: alanine aminotransferase; BSA: body surface area; BMI: body mass index; BUN: blood urea nitrogen; SCr: serum creatinine; eGFR: the estimated glomerular filtration rate; HCT: red blood cell specific volume; HGB: hemoglobin; HT: height; LTG: lamotrigine; LEV: levetiracetam; VPA: co-administration with valproate acid; WT: weight.

It was found that eGFR significantly affected MHD CL, according to the observations from Lin et al. [29]. In patients with impaired renal function (eGFR: 20–80 mL/min), the CL was 34.8% (9.4%–64.2%) lower than those with normal renal function.

There was one study identified dose could affect CL, which may indicate a nonlinear elimination of MHD [27]. However, these effects may on account of the dose adjustment in clinical practice according to the TDM process, which can be misinterpreted as nonlinearity in the system [37]. Therefore, further in-depth study on the possible PK of MHD is necessary.

The influence of all included covariates on CL is shown in **Fig. 4**. The ranges of continuous covariates that were scaled are as follows: the weight of adults was set at 40–100 kg and that of children was set at 16–40 kg. The age was set as 18–75 years. The range of eGFR and alanine aminotransferase (ALT) were set as 20–120 mL/min and 5–150 U/L, respectively. The dose of OXC was set as 75–1350 mg/kg/d.

**Fig. 4.**
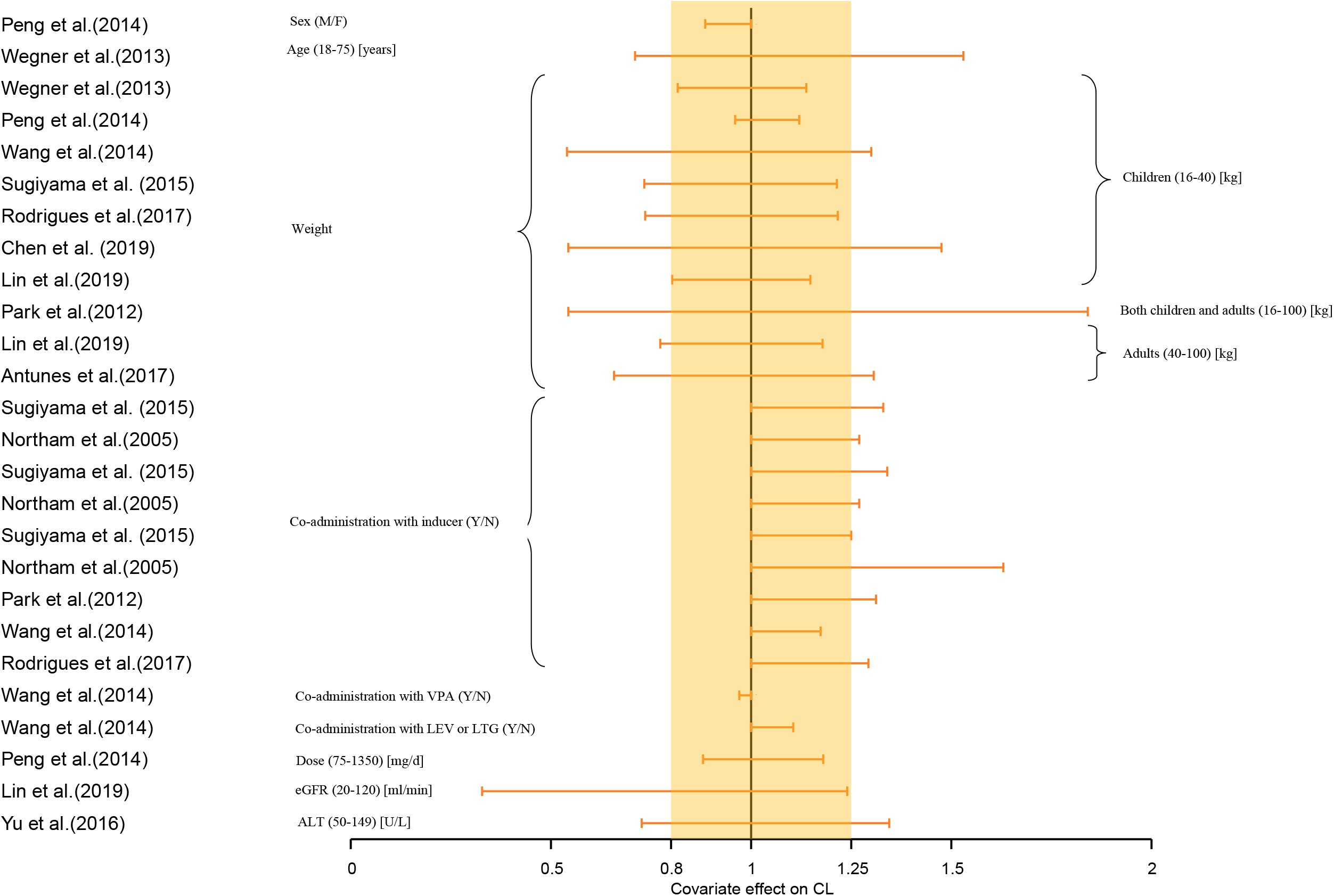
Covariate effect on the clearance of oxcarbazepine. The horizontal bars represent the covariate effect on clearance in each study. The typical value of clearance in each study was considered to be 1. The effect of each covariate for clearance is displayed by the ratio of clearance in the range of each covariate to the typical clearance value. The shaded gray area ranges from 0.8 to 1.25. ALT: alanine aminotransferase; eGFR: the estimated glomerular filtration rate; LEV: levetiracetam; LTG: lamotrigine; VPA: co-administration with valproate acid; Y: yes, co-administration; N: no, not co-administration; M: male; F: female.

Ten of all studies that investigated impact of the weight indicated that it was associated with the CL of MHD [24,26-32,34,35]. Weight had a significant influence on CL. Our finding showed that the CL of patients with different weight could range from 0.54 to 1.8 times compared to the typical patient (children: 30kg, adults: 70kg). Co-administration with EIAEDs also significantly affected CL in children and adults, which approximately increased 1.17 to 1.63 times [24-26,28,32]; in four studies, the clinical significance of the impact of EIAED co-administration on CL was more than 20% [24-26,32]. eGFR was identified to significantly affect the CL of adults with a range of 0.33 to 1.24 times [29] compared to a typical patient with an eGFR of 90 mL/min. ALT was also found to have a significant impact on CL in children, with a range of 0.73 to 1.35 times compared to a typical patient with an ALT level of 30 U/L [33].

Four studies conducted a model-based simulation to show the influence of covariates on the PK profile and optimize the dosage regimen to achieve the target MHD concentrations of 3 and 35 mg/L recommended by the current guideline [29-32]. For adults and paediatric patients, all the four studies recommended adjusting the dosage [29-32]. Chen et al. [31] and Rodrigues et al. [32] had a similar recommended dose range for paediatric patients based on their weight. However, Lin et al. [30] recommended a lower dose than others, which may be explained by the lower CL in their study than others (0.04 L/h/kg vs. 0.059/0.057 L/h/kg). Meanwhile, Rodrigues et al. [32] also recommended increasing the maintenance dose by 50% above the recommended maintenance dose for children treated with EIAEDs.

## 4. Discussion

To our knowledge, this is the first review that summarised data concerning previously published PPK models of OXC and its active metabolite MHD. We found considerable PK differences between children and adults. We also provided evidence regarding adjustment of the dose regimen according to weight, co-administration with EIAEDs, and eGFR.

The PK of MHD showed no significant differences due to ethnicity. According to the PPK studies, there were similar PK profiles in adults. The weight-standardised CL in Caucasians (median: 0.045, range: 0.029–0.060 L/h/kg) was similar to that in Asians (median: 0.036, range: 0.033–0.039 L/h/kg) (P > 0.05). Likewise, most PPK studies performed in paediatric patients also displayed comparable PK profiles. Moreover, Sugiyama et al. [24] directly compared the PK of MHD among Caucasian, Black, and Asian populations and found no obvious differences among these sub-populations. Nevertheless, studies conducted in Chinese populations showed larger variabilities than other ethnicities (0.016–0.084 L/h/kg vs. 0.043–0.078 L/h/kg). The reason for this is unclear and may require still exploration.

The PK of MHD displayed considerable difference among infant, children, and adult patients. Adults generally displayed lower CL per kg than the infants and children, consistent with the classical PK investigations [38]. This difference may be attributed to the lower body fat/lean mass ratio, higher kidney blood flow, and higher total body water in children [39]. Furthermore, children had a wider weight-standardised CL range than adults, which is attributed to the maturity of the liver and inefficient UGT enzyme capacity [40,41]. A higher CL resulted in a lower concentration of MHD. Therefore, a higher dosage could be used in paediatric patients to achieve the target steady-state concentration of MHD [23].

Recently, OXC was increasingly off-label used to treat epileptic seizures in infants [39] owing to its linear PK, better safety, and well-tolerated profile [6]. Piña-Garza et al. [42] reported that OXC is relatively safe and effective in the treatment of infant seizures. To date, only one PPK study has been conducted in paediatric patients aged 0.2 to 3.75 years [25]; therefore, clinical data of OXC in the treatment of infants are limited. To ensure better efficacy and safety in infants, further studies are necessary to explore the covariates that affect the variability of OXC.

Previous PPK studies found that EIAEDs could significantly increase the CL of MHD. This is consistent with the PK study performed by McKee et al. [43], who found that CBZ and phenytoin (PHT) decreased the AUC of MHD by 40% and 29%, respectively. As MHD is primarily cleared via glucuronic acid conjugation, EIAEDs may play a more important role through the induction of UGT-mediated glucuronidation to decrease serum MHD concentration [35]. Lower exposure could increase the risk of seizures [44]; therefore, the dose of OXC may be increased when co-administered with EIAEDs.

Renal function may also influence the CL of MHD because OXC and MHD are almost completely excreted by the kidney (94%–97.7%) [5]. Lin et al. [29] found that MHD CL decreased by 64.2% when the eGFR decreased to 20 mL/min and recommended dose adjustment according to eGFR in adult patients. When the eGFR is normal, at approximately 90–120 mL/min, the recommended dose is 225 or 300 mg q12h [29].When the eGFR is less than 30 mL/min, the recommended dose is 75 mg q12h. This was similar to the OXC label recommendation that indicated that patients with impaired renal function (creatinine clearance <30 mL/min) should initiate OXC at one-half of the dose of 300 mg/day q12h. Therefore, to ensure a safe dose for adults with renal dysfunction, a more detailed dose recommendation may be required.

However, there are also PPK studies that did not identify eGFR as a covariate [24,33]. This may be because of the low proportion of study cohorts with low eGFR levels. Thus, patients with renal impairment on MHD clearance warrant further investigation to optimise their dosing regimen.

There were many unknown variabilities in children that could not be explained by previously reported PPK models. In the included studies, children had a larger RUV (additive model 0.93–5.12 mg/L; proportional model 4%–32%) than adults (RUV: additive model 2.38–3.80 mg/L; proportional model 1.7%–24.5%). Moreover, the practice guideline for the TDM of AEDs indicated the benefits of TDM for children such as diagnosis of clinical toxicity and guide dosage adjustment [45]. Thus, TDM monitoring is more crucial for paediatric patients taking OXC.

## 5. Conclusions

The PPK studies of OXC have been extensively reviewed. The PK of MHD has a large difference between children and adults. Furthermore, children have a larger variability in PK than adults. This review showed that to optimise the dosing regimen of OXC, the weight, identity of co-administered drugs, and renal function of patients should be considered.

## 6. Expert Opinion

Long-term maintenance therapy with drugs is one of the important ways of anti-epilepsy. It is high requirements for maintenance dose and dose adjustment, and most antiepileptic drugs have great variability. Therefore, in order to improve the safety and effectiveness, researchers have conducted PPK studies on antiepileptic drugs, hoping to find the potential covariates that affect its PK variation.

Although, there are many PPK models established, few articles summarised the information of the published PPK studies. This review attempts to summarize the knowledge concerning the PPK modeling of oxcarbazepine to determine the factors that can affect its clearance in the body. We find that there are no significantly difference on races. However, our research shows that children have more variation than adults. Finally, we also explored the significant covariates that may have clinical impact to provide evidence for adjusting dosage regimens of oxcarbazepine.

Owing to the better PK profile and formula diversity, it is increasingly used in the special populations, such as infants and elders. Infants have great variability because of their physiological characteristics and TDM may help to optimize the dosage regimen. However, most hospitals do not realize the importance of TDM monitoring. In the future five years, it is necessary to conduct further studies on infants to find significant covariates that affects their PK variation. Nevertheless, the covariates selected are often different from the theory. This may be due to the smaller variation range of the covariates or the lower proportion of the covariates with large variation range. Therefore, in the future PPK studies, we need to focus on the collection of people with large variation of potential covariates suspected in clinical practice. We maybe get objective and credible results.

In clinical practice, we need to optimize dosing according to PK/PD. PPK studies had shown that the co-administration with inducers can increase the dose of OXC to achieve the therapeutic target concentration. However, there is no corresponding PD index to determine whether the dose of OXC should be increased in combination with inducers. Therefore, it is very important to establish PK / PD model to explore the relationship between dose-exposure-response. Furthermore, we can also use the established PK / PD model to refine the treatment window in the different disease severity and population.

### 6.1 Five-year Review

In order to optimize the clinical administration scheme and reduce the occurrence of adverse events, PPK modeling was conducted for drugs with large variability and narrow treatment window. Although many models have been established, there is no unified standard for the quality of effective identification models. External validation is the most. In the five future years, it will explore the criteria for evaluating the quality of models and apply them to evaluate the quality of published models.

The ultimate purpose of establishing PPK model is to solve practical difficulties. However, the application of PPK models is mainly based on dose simulation. In the future, exploring meaningful clinical application is one of the main development directions of PPK study. For instance, the PPK model is used for the remedial administration of missed late administration. Epilepsy patients are stricter with the time of taking drugs. Once the occurrence of missing a dose, it is likely to relapse epilepsy. FDA guidelines for remedial administration of antiepileptic drugs are vague. Therefore, PPK models plus TDM can be used to give more precise and quantitative remedy for patients with missed dose. It can minimize the deviation time from the treatment window and maximize the benefit of patients.

## Data Availability

Not applicable

## Acknowledgments

The authors would like to sincerely thank *Dr. Vincent Jullien* from Service de Pharmacologie, Hôpital européen Georges Pompidou, France; *Yang Wang* and *Jing Peng* from Department of Pharmacy, Wuhan Children’s Hospital, China; *Yunli Yu* from Department of Clinical Pharmacology, The Second Affiliated Hospital of Soochow University, China for providing details about the research and active discussions on the coding. We would like to thank *Hai-ni Wen* MPharm from Shanghai Chest Hospital; and Ph.D. candidate *Xiao-qin Liu* from Department of Pharmacy, Huashan Hospital, Fudan University, China for their critical comments. We would also like to thank Editage (www.editage.cn) for English language editing.

## Funding

Not applicable.

## Declaration of interest

Yue-ting Chen, Chen-yu Wang, Yi-wei Yin, Zi-ran Li, Wei-wei Lin, Min Zhu and Zheng Jiao declare that they have no conflict of interest.

## Author contributions

**Yue-ting Chen:** Data curation, Writing-Original draft preparation, Writing-Reviewing and Editing, Visualization; **Chen-yu Wang:** Data curation, Writing-Reviewing and Editing; **Yi-Wei Yin:** Writing-Reviewing and Editing, Supervision; **Zi-ran Li:** Software, Writing-Reviewing and Editing; **Wei-wei Lin:** Software, Validation, Writing-Reviewing and Editing; **Min Zhu:** Software; **Zheng Jiao:** Conceptualization, Methodology, Writing-Reviewing and Editing, Supervision.

## Notes

### Competing Interest Statement

The authors have declared no competing interest.

