## Supplemental Table 1 for "Population Pharmacokinetics of Oxcarbazepine: A Systematic Review"

**Supplementary** **material**

**Table S1. List of tested and significant covariates in the models.**

| **Study**  **(publication year)** | **Tested covariates** | | | **Covariate selection criteria** | | **Significant covariates** | | |
| --- | --- | --- | --- | --- | --- | --- | --- | --- |
|  | **Demographic** | **Laboratory tests** | **Co-administration** | **Forward inclusion** | **Backward elimination** | ***CL*** | ***V*** | ***Tlag*** |
| Northam et al. (2005) | BSA^a^, HT | NR | CBZ, PB, PHT | NR | NR | BSA^a^, CBZ, PB, PHT | HT | NR |
| Park et al.  (2012) | Age, WT, Sex | NR | EIAEDs^b^、  non-EIAEDs^c^ | P < 0.05 | P < 0.01 | WT, EIAEDs^b^ | NR | NR |
| Wegner et al. (2013) | Age, WT, Sex | NR | NR | P < 0.01 | NR | WT, Age | NR | NR |
| Wang et al.  (2014) | Age, WT, Sex, BSA | NR | EIAEDs^b^, NEWAEDs^d^, VPA | P < 0.05 | P < 0.01 | EIAEDs^b^, WT, NEWAEDs^d^, VPA | WT | NR |
| Peng et al.  (2014) | Age, WT, Sex, Dose | NR | VPA, CBZ, CZP, NTZ, PB, TPM, LEV, LTG | P < 0.05 | P < 0.01 | Sex, Dose, WT | Age, WT | NR |
| Sugiyama et al. (2015) | WT, BSA, Race | Cr, AST, ALT | CBZ, PB, PHT | P < 0.05 | P < 0.01 | WT, CBZ, PB, PHT | WT | NR |
| Yu et al.  (2016) | Age, Sex | AST, ALT, BUN, Cr, RBC, HGB, HCT | NR | P < 0.05 | P < 0.01 | ALT, Sex | BUN, HGB | Age |
| Antunes et al. (2017) | WT | NR | verapamil | NR | NR | CL total: WT, CL(MHD): WT | Vc, Vp, V(R-MHD), V(S-MHD): WT | NR |
| Rodrigues et al. (2017) | Age, WT, Sex | NR | EIAEDs^b^ | P < 0.05 | P < 0.01 | CL(OXC): WT  CL(MHD): WT, EIAEDS | Vc(OXC): WT  Vc(MHD): WT | NR |
| Chen et al.  (2019) | Age, WT, Sex, HT, BSA, BMI | HGB, HCT, ALT, AST, TP, ALB, Cr, Blood Sodium, BUN | EIAEDs^b^, VPA, NEWAEDs^d^ | P < 0.05 | P < 0.01 | WT | WT | NR |
| Lin et al.  (2019) | Age, WT, Sex,  [genetype](javascript:;) | ALT, AST, Cr, eGFR^e^, BUN | VPA, NEWAEDs^d^ | P < 0.05 | P < 0.01 | WT | NR | NR |
| Lin et al.  (2019) | Age, WT, Sex,  Genetic factors | ALT, AST, Cr, eGFR^e^, BUN | VPA, NEWAEDs^d^ | P < 0.05 | P < 0.01 | WT, eGFR^e^ | NR | NR |

ALT: Alanine Aminotransferase; AST: Aspartate Aminotransferase; BSA: body surface area; BUN: Blood urea nitrogen; CBZ: carbamazepine; Cr: serum creatinine; CZP: clonazepam; DDPW: Daily dose per weight; EIAEDs: Enzyme-inducing antiepileptic drug; GPB: gabapentin; HCT: Red blood cell specific volume; HGB: Hemoglobin; HT: Height; LEV: levetiracetam; LTG: lamotrigine; NEWAEDs: newer antiepileptic drug; NTZ: nitrazepam; PB: phenobarbital; PHT: phenytoin; RBC: Red blood cell counts; TPM: topiramate; TP: total protein; VPA: valproic acid; WT: Weight.

^a^BSA=$0.023\times{(HT)}^{0.422}\times{(WT)}^{0.515}$;

^b^EIAEDs: carbamazepine, phenobarbital, phenytoin;

^c^non-EIAEDs: lamotrigine, topiramate, zonisamide, pregabalin, levetiracetam, clobazam, clonazepam, and gabapentin;

^d^NEWAEDs: levetiracetam, lamotrigine, topiramate;

^e^eGFR was calculated by the CKD-EPI equation.
